# Automated and impact-based quality control in multiple-breath washout

**DOI:** 10.1101/2025.09.23.25336339

**Authors:** Florian Wyler, Silvio Borer, Marion Curdy, Xenia Bovermann, Bettina Frauchiger, Philipp Latzin

**Affiliations:** Division of Paediatric Respiratory Medicine and Allergology, Department of Paediatrics, Inselspital, Bern University Hospital, University of Bern, Bern, Switzerland; Graduate School for Health Sciences, University of Bern, Bern, Switzerland

**Author notes:** **Correspondence** Philipp Latzin, Division of Paediatric Respiratory Medicine and Allergology, Department of Paediatrics, Inselspital, Bern University Hospital, University of Bern, Freiburgstrasse 8, Bern 3010, Switzerland.

## Abstract

**Introduction:** Quality control (QC) is an important but challenging step in the correct interpretation of multiple-breath washout (MBW) tests, as irregular behaviors can lead to biased outcomes.

**Methods:** We developed an automated QC algorithm to produce estimates of the impact of irregular behaviors on MBW outcomes. It compares results between simulated measurements containing either ideal target behaviors and the observed irregular behaviors of the measurement. Differences in main MBW outcomes (Lung clearance index (LCI), functional residual capacity) between simulations served as behavior-specific QC outcomes. We applied the automated QC algorithm to 2471 measurements of 87 children with cystic fibrosis (CF), and compared the automated QC with that performed by experienced raters (expert QC) in a dataset of 100 measurements of healthy children and children with CF.

**Results:** The test could be applied successfully to all measurements. We found that the impact of QC-related factors on MBW outcomes explained around 45% of the within-visit variability of LCI in children with CF. The overlap between automated and expert QC was moderate but comparable to the overlap between two separate instances of expert QC. The factors with the largest effect on MBW outcomes were found to be irregular increases in expired N2 concentration (leaks, trapped gas, in 31% of cases), changes in end-expiratory lung volume (23%) and variations in breath size around the end-of-test (19%).

**Discussion:** We developed a novel automated QC tool to produce estimates for the accuracy of MBW outcomes, providing a fast, reproducible and impact-based method to perform QC for MBW measurements.

## Introduction

Multiple-breath washout (MBW) is an established technique to assess ventilation inhomogeneity in the lungs^1–4^. In nitrogen (N_2_) MBW, subjects wash out the resident N_2_ in their lungs using pure oxygen (O_2_). The test’s main outcomes are the lung clearance index (LCI) and the functional residual capacity (FRC). MBW tests typically last several minutes, and while they mostly require only passive cooperation from the test subject, there are irregular behaviors (e.g. irregular or non-tidal breathing, coughs, leaks), which can lead to biased or unusable test outcomes ^5–8^. A quality control (QC) step, which identifies such irregular behaviors, is therefore an important component of the correct interpretation of MBW measurements. The application of QC has been shown to have a significant impact on the variability of MBW outcomes, particularly on repeatability of LCI.^9–12^.

There are existing QC guidelines for MBW, which allow experts to make judgments on whether they should include or exclude measurements for research and clinical routine ^13–15^. These guidelines allow expert raters (typically trained physicians, nurses or research scientists) to assess whether the test subject behaved in a way which led to an adverse effect on the test’s main outcomes, independent of the ventilation inhomogeneity that is being measured.

Performing this expert QC on MBW data represents the current gold standard, but it has several major limitations:

i. Expert QC is very time-consuming and the level of expertise required for it is not common in clinical routine. This represents a barrier for more widespread use of MBW.
ii. In cases where the irregular behavior is of intermediate quality - between clearly acceptable and clearly unusable - expert QC has a significant subjective component that might lead some raters to accept and others to reject a given measurement. This is especially the case when there are multiple instances of minor irregularities within one measurement. As MBW measurements are typically several minutes long, and as they are also performed in awake people starting in pre-school age, it is very common for them to contain such “grey zone” irregular breathing, especially if a lung pathology is involved. This subjectivity of QC, outside of dedicated central over-reading centers for MBW data, makes consistent QC difficult and potentially leads to non-reproducible results with a difficult-to-quantify bias.
iii. While it is feasible to spot irregularities in the signals of a MBW measurement, it is very challenging to estimate the impact or importance of such irregularities on LCI or FRC. Expert QC therefore relies on empirically determined thresholds for allowed variations in behavior for each individual test.

There is therefore a need for tools to automatically perform MBW-QC in an objective, reproducible manner.

## Methods

### Study design

This is a proof-of-concept study to i) develop a novel quality control (QC) tool and ii) assess its practicability using existing datasets of healthy children and children with cystic fibrosis (CF). The study was approved by the ethics committee of the Canton of Bern (2019-01072, 2017-02139).

### Development of the QC tool

#### Objective QC

In order to develop an objective automated QC algorithm, we aimed to relate potentially problematic breathing irregularity events (e.g. coughs, non-tidal breathing, leaks) directly with estimated changes in multiple-breath washout (MBW) outcomes. We therefore developed a model, which - for any given breath - related an input of breathing behavior with an output of expected Nitrogen (N_2_) dilution. To do this, it took into account the current end-expiratory lung volume (EELV), physiological and machine dead spaces, and how the N_2_ washout efficiency changed over the course of washout for a given test subject.

Using this model we ran washout simulations to make side-by-side comparisons between simulated measurements with a given observed irregularity (e.g. the observed changes in EELV during a measurement) and simulated measurements without said irregularity (i.e. if EELV had remained constant). If the two simulations finished with similar outcomes, we could conclude that the irregularities in question were minor enough not to affect main outcomes. However, if one measurement finished significantly sooner or later than the other, we could conclude that the irregularity in question was expected to have a significant effect on MBW outcomes.

#### MBW model

The washout model we used to simulate measurements consisted of the following elements: A mixing alveolar volume (V_A_), a non-mixing volume of dead space (V_D_), and a decreasing dilution efficiency as a function of N_2_ washed out of the lungs. The input to this model was a series of ingoing and outgoing breath volumes (V_insp_, V_exp_). The dilution of N_2_ was modelled to take place in the ventilated volume V_A_. Before inspiration, the N_2_ was contained in the volume V_A_ + V_D_, and after inspiration it was contained in volume V_A_ + V_D_ + (V_insp_ – V_D_) = V_A_ + V_insp._ Therefore, the dilution factor K was equal to K= (V_A_+V_D_)/(V_A_+V_insp_) (Figure 1). This dilution was modelled to be modified by an efficiency factor (f), which either stayed constant or decreased over the course of the washout as a function of how much N_2_ was left in the lungs (supplementary figure S1). The changing efficiency of this dilution simulated the fact that well-ventilated compartments initially wash out more efficiently in MBW, and that over time most of the remaining N_2_ is situated in less well-ventilated parts of the lungs, slowing down the washout process^16^. An efficiency factor f=1 in the washout model corresponded to *homogenously ventilated lungs*.

**Figure 1:**
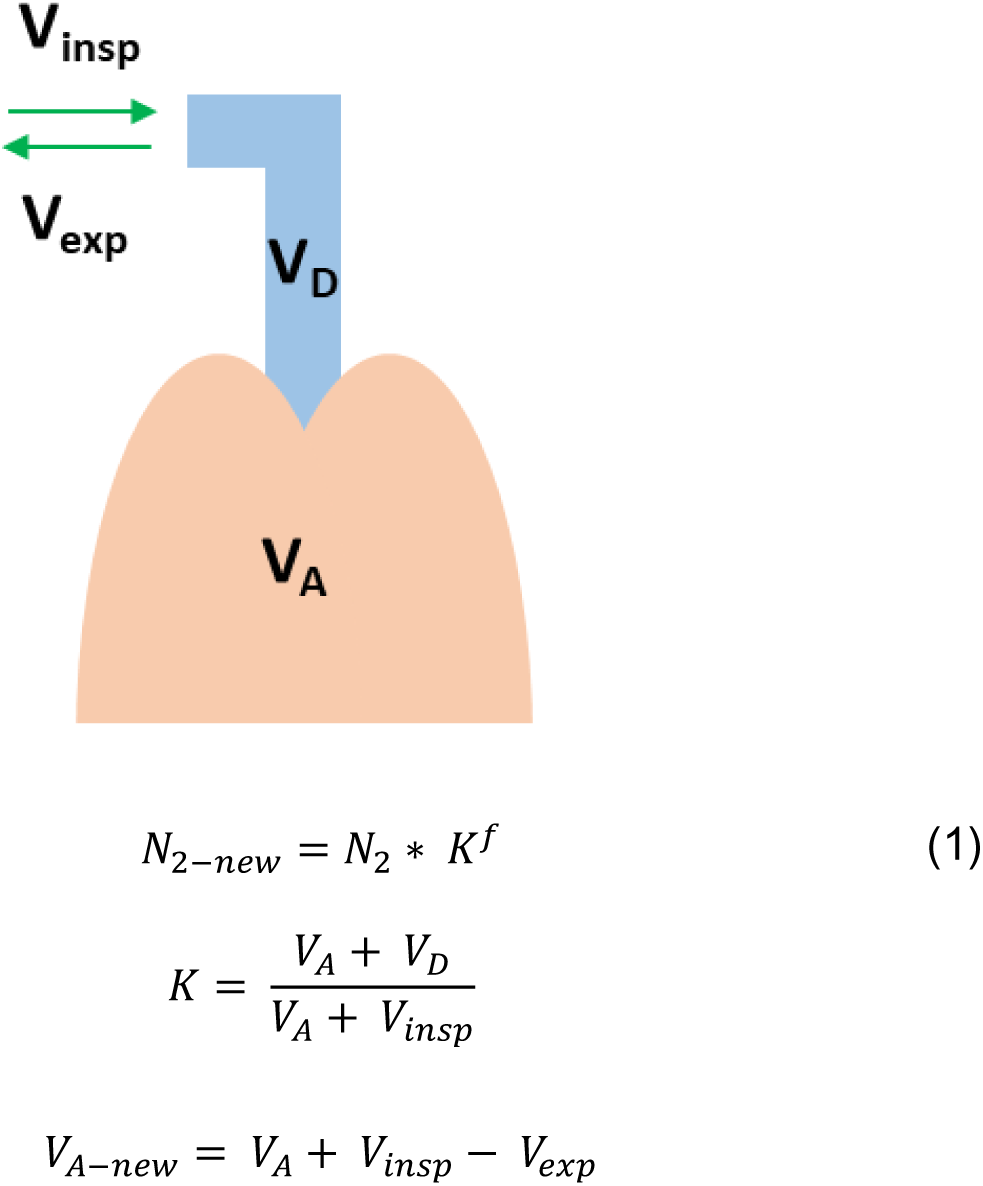
Illustration of the washout model. The input to the model is a series of inspired and expired volumes (V_insp_, V_exp_). The inspired volume that traverses the dead space V_D_ equals (V_insp_-V_D_) and dilutes the concentration of N_2_ within volume V_A_, resulting in N_2-new_. The difference between V_insp_ and V_exp_ modifies the alveolar volume to V_A-new_ for the next breath. As the washout progresses and N_2_ washes out, the efficiency f of the dilution decreases.

We defined the washout progress of a breath (P_breath_) as the difference between normalized logarithmic end-tidal concentrations (supplementary equation S2) of N_2_ between two breaths. Expressing washout progress this way allowed a more direct comparison between dilutions throughout the washout. A breath of the same size and efficiency leads to different changes in absolute end-tidal concentrations at the start and end of washout. However, the same dilution of N_2_ occurs, and therefore the same progress P_breath_. Normalization was chosen such that the cumulative washout progress (P_cumulative_) required to complete an MBW measurement was equal to 1. Consequently, ten breaths with P_breath_ = 0.1 were expected to finish a measurement, equivalent to five breaths with P_breath_ = 0.2. We defined as washout efficiency f of a breath the ratio between the measured progress of a breath and the progress predicted by the MBW model assuming homogenously ventilated lungs.

The parameters used by the MBW model (V_A_, V_D_, and efficiency factor f(P_cumulative_)) were estimated from each MBW measurement individually. V_D_ could be estimated from the CO_2_ and flow signals (by measuring at which expired volume there was a rise in CO_2_), and V_A_ was equal to FRC minus V_D_. Efficiency was first calculated per breath by comparing the observed dilution of N_2_ with the predicted dilution in homogenously ventilated lungs. Second, the resulting efficiency values per breath were used to determine a linear model of efficiency as a function of cumulative washout progress, excluding outliers (Supplementary Figure S1). This linear fit could be used to determine the efficiency factor for a given breath as a function of the washout progress.

#### Quality control tests

Current QC guidelines focus on identifying behaviors which are known to impact LCI and/or make it less meaningful. These factors include:

i. breathing artifacts such as changes in EELV, breaths of the wrong size (especially at the beginning or end of test), hypo-or hyperventilation,
ii. leaks or trapped gas released throughout the measurement, leading to unexpected rises in end-tidal N_2_, or
iii. technical issues such as breathing artifacts which might lead to early termination of the measurement, insufficient wait time between measurements and signal synchronization issues.

We developed a set of tests for automatic quality control. For each test we chose a potential source of error from the above list (either behavioral or methodological), and attempted to estimate its effect by one of two ways:

i. running two simulations using the washout model, where the inputs differed by the behavior in question, or
ii. analyzing the measurement with two differing methods (e.g. using different breath detection or signal synchronization methods).

Each test resulted in a*n estimated effect,* defined as the difference between the results obtained with a given behavior or methodology and those without the behavior or with an alternative methodology (expressed in percentage).

These tests, together with simple numerical tests for initial N_2_ concentration and inspiratory overflow, were chosen to cover the same irregular behaviors and sources of error as those examined in current expert QC procedures. All tests can be evaluated in an automated, reproducible manner. The specific tests are listed below.

#### End-expiratory lung volume (EELV)

In order to test for the effect of changing EELV, we compared the MBW outcomes of i) washout simulations using the measured V_insp_ and V_exp_ of the measurement as input, vs. ii) washout simulations using the measured V_insp_ but setting V_exp_ = V_insp_, which leads to a stable EELV over the course of the measurement.

#### Tidal volume (VT) variability

To test for the effect of variable tidal volumes (V_T_), we compared i) washout simulations using measured V_insp,_ but where V_exp_ = V_insp_ (resulting in stable EELV but variable V_T_), vs. ii) washout simulations where all V_insp_ and V_exp_ were equal to the median V_T_ of the measurement (resulting in stable EELV and stable V_T_).

#### VT size

To test for the average effect of the measured V_T_ size, we compared i) washout simulations where all V_insp_ and V_exp_ were equal to the median V_T_ of the measurement vs. ii) washout simulations where all V_insp_ and V_exp_ were equal to the most efficient V_T_, calculated based on estimated dead-space V_D_ and alveolar volume V_A_ (see OLS section 1.3).

#### Leak / Trapped gas

To test for the impact of unexpected upticks of N_2_ concentration (due to either leaks or trapped gas released throughout the measurement), we compared i) washout simulations where the end-tidal concentration of N_2_ was equal to the measured N_2_ vs. ii) washout simulations where end-tidal N_2_ was simulated by the washout model. As the washout model ignored major outliers when fitting efficiency to washout progress, the washout simulation provided a reference for an average washout and would not incorporate leaks.

#### End-tidal concentration measurement

To test for the effect of breath size on end-tidal concentration and consequently end-of-test determination, we compared the outcomes of i) a standard MBW analysis vs. ii) a more robust MBW analysis with an improved size-independent method for determining the end-tidal concentration (see OLS section 1.4).

#### Synchronization

To test for the effect of calibration on the signal synchronization we compared the outcomes of i) a standard MBW analysis vs. ii) an MBW analysis with recalculated synchronization (see OLS section 1.5).

#### Breath detection

To test for the effect of small breaths and irregular breathing patterns which could be misinterpreted as an early termination of the measurement, we compared the outcomes of i) a standard MBW analysis vs. ii) a more robust MBW analysis with a breath detection that did not remove breaths and which had a dead-space V_D_-dependent breath detection threshold^17^.

### Application and Validation of the QC tool

#### Datasets

To characterize the results of automated QC and the effect of irregular behaviors on measured within-visit variability, we re-analyzed an existing dataset of 4490 MBW measurements, originating from 1618 longitudinal visits of 87 children with CF. The measurements were conducted between 2012 and 2019. A smaller version of this dataset has been described in detail elsewhere^18^. In order to make within-visit correlations of QC-related estimates with MBW outcomes, we included only visits which had at least three completed measurements.

To compare the outcomes of automated QC with QC performed by expert raters (expert QC), we randomly selected 50 completed MBW measurements each from 6 year old healthy children and 6 year old children with CF from measurements conducted between 2020 - 2025 in the Bern Basel Infant Lung Development (BILD)^19^ and Swiss Cystic Fibrosis Infant Development (SCILD)^20^ cohorts respectively. This dataset was chosen to be a representative sample of measurements from clinical routine, in an age group most likely to produce QC-relevant behaviors.

Nitrogen (N_2_) MBW measurements were collected using the Exhalyzer® D (Ecomedics AG, Duernten, Switzerland), and analyzed using Spiroware 3.3.1.

#### Descriptive application

To characterize the application of the QC tool to the real datasets of healthy children and children with cystic fibrosis, we determined feasibility, time requirements, and the QC tests with the largest and most frequent impact on MBW outcomes

#### Association to LCI

Each of the quality control tests provided an estimate of the impact of individual irregularities on MBW outcomes. Most tests estimated an effect on cumulative expired volume (CEV), i.e. the length of the test, while others (synchronization, leaks) estimated both an effect on CEV as well as on FRC. To obtain a single impact estimate of the effect on LCI, we multiplied the individual estimated effects on CEV originating from the different QC tests, and divided this by the effects on FRC.

In order to compare within-visit variability of LCI with the QC-based impact estimate across visits and individuals, we divided both the LCI and estimated impact on LCI by the mean value of the given visit.

#### Comparison to expert QC

The automated QC algorithm produced an estimate for the impact of QC-related factors on MBW outcomes. In order to obtain a single binary outcome of acceptability per measurement, a threshold of acceptable estimated impact had to be defined. This threshold can be adapted depending on the severity of QC desired. We defined measurements to have acceptable QC if no individual test had a higher QC-related change in MBW outcomes than 15%, and the numerical thresholds (e.g. for initial concentration) were passed.

In order to compare the outcomes of automated QC with expert QC, we compared the binary outcome of “accepted/rejected” from the automated QC with the original QC performed by the expert rater conducting the measurement, as well as a second round of retrospective QC performed by a different experienced expert rater.

#### Statistics

To compare automated QC with one of the two expert QC, we computed Cohen’s kappa between each pair of the three instances of QC.

To evaluate the association between relative LCI and aggregated or individual QC impact estimates, we computed coefficients of determination (R^2^).

The agreement of within-visit variability of LCI with estimated QC impact was measured by computing the Pearson correlation coefficient for the relative measurement outcomes of each visit.

Statistics were performed using MATLAB R2022a (The MathWorks Inc., Natick, Massachusetts, USA), and Python 3.12 (Python Software Foundation).

## Results

### Datasets

In the large dataset of 87 children with cystic fibrosis (CF, dataset 1), a total of 2471 measurements from 754 visits by 84 participants had at least three measurements per visit, and were included in the analysis (Table 1).

**Table 1:** Participant demographics for dataset 1 (composed of children with CF aged 3-18 years), and dataset 2 (composed of randomly sampled measurements of 6 year-old children with CF and HC). For dataset 1 only visits with at least three completed measurements were included. Descriptive statistics are median (minimum, maximum), and are computed across all visits, and across all individual measurements for LCI, FRC. Abbreviations: CF, cystic fibrosis; HC, healthy control; LCI, Lung clearance index; TO, turnover; FRC, functional residual capacity.

| DATASET 1 - CF |  | DATASET 2 – CF / HC |  |
| --- | --- | --- | --- |
|  | CF | CF | HC |
| SUBJECTS (N) | 84 | 22 | 40 |
| VISITS (N) | 754 | * | * |
| MEASUREMENTS (N) | 2471 | 50 | 50 |
| FEMALE: N (%) | 52 (62) | 12 (55) | 19 (48) |
| All Visits |  |  |  |
| AGE (YEARS) | 10.5 (3.9, 18.9) | 6.2 (6.0, 6.6) | 6.1 (6.0, 6.5) |
| WEIGHT (KG) | 30.8 (15.7, 80.7) | 22.1 (15.7, 30.7) | 23.5 (17.8, 38.4) |
| HEIGHT (CM)= | 137 (100, 185) | 118 (102, 138) | 119 (106, 147) |
| Individual measurements |  |  |  |
| LCI 2.5% (TO) | 8.4 (3.1, 18.7) | 7.2 (5.5, 10.6) | 6.1 (5.1, 8.1) |
| FRC (L) | 1.1 (0.4, 5.4) | 0.9 (0.6, 1.6) | 1.0 (0.6, 1.6) |

The smaller dataset of randomly selected measurements from 6-year-old children included 50 measurements from 22 children with CF, and 50 measurements from 40 healthy children (Table 1).

### Objective QC

It was possible to apply the quality control algorithm to all measurements in a fully automated manner, without any manual intervention. Evaluation time of a single measurement (including MBW analysis) on an average office desktop computer was on the order of 4 seconds using the current non-optimized implementation of the automated QC code, which translates to 7 minutes for 100 files.

### Quality control tests

An illustrative example of the automated QC method using the lung model is shown in Figure 2. It shows the comparison between a simulation using the measured breathing volumes of a measurement (including breathing irregularities, here specifically changing EELV) and the resulting washout progress, compared to one without EELV irregularity, in two different MBW measurements. For the two cases the washout progress differs resulting in different measurement end points and thus MBW outcomes.

**Figure 2:**
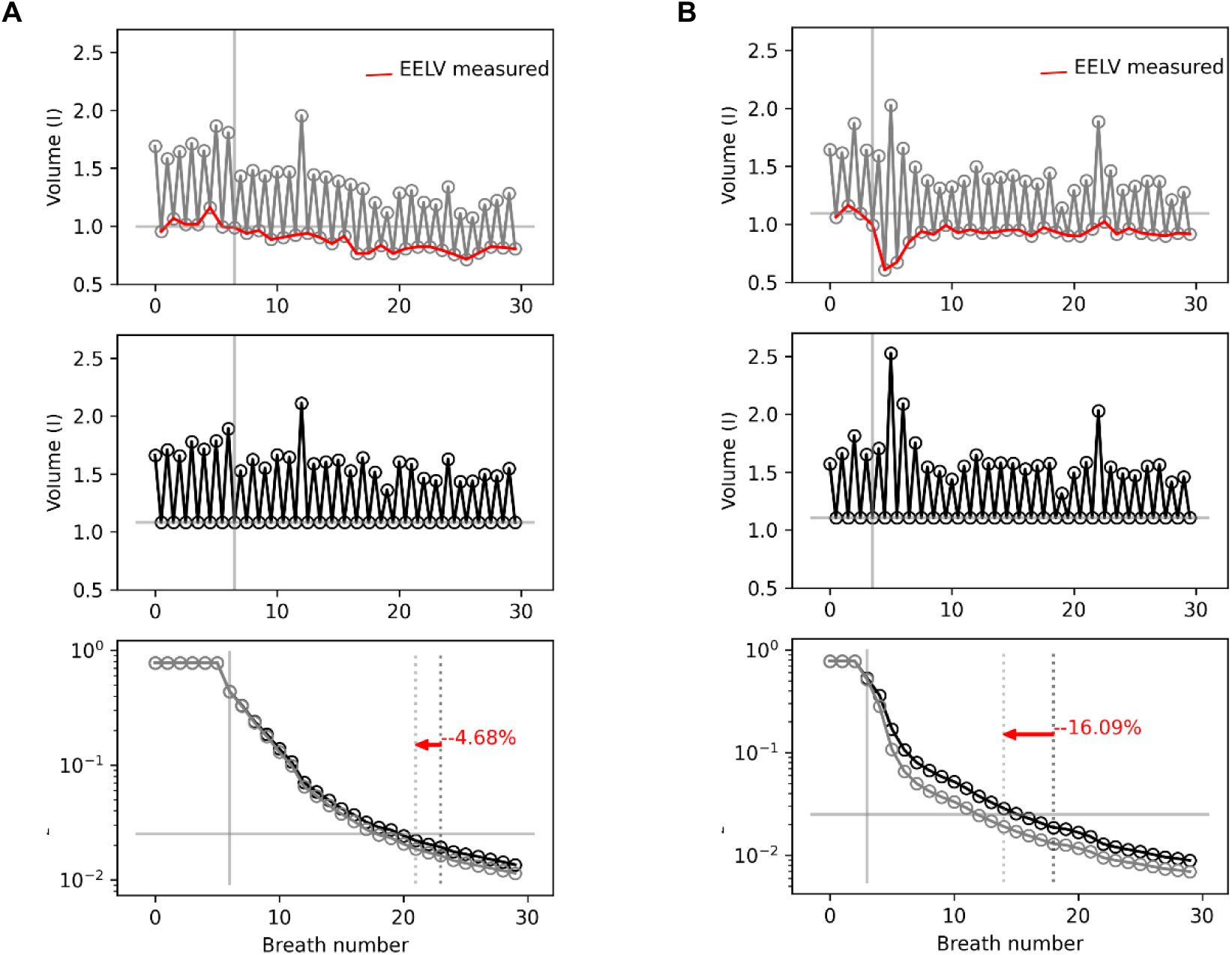
Illustration of the EELV QC test for measurements of one example visit. Panel **A** illustrates the EELV test for an accepted, Panel **B** for a rejected measurement by the EELV test criterion. **A, B** Top: inspiration and expiration volumes are shown per breath. Middle: To test for influence of variation in EELV the expiration volume is set equal to inspiration volume. This results in a steady EELV over the washout. The horizontal line indicates the calculated FRC. Bottom: Washout simulated with the washout model using the true volumes of top panels resulting in the N_2_ concentration shown in grey. The resulting N_2_ concentration of a simulation using steady EELV (middle panel) is shown in black. The vertical solid grey line shows the first washout breath, the vertical dotted lines show the washout end for each simulation. The horizontal grey line shows the threshold N_2_ concentration (2.5%). The EELV test estimates an effect of changing EELV on LCI of −4.7% for the measurement shown in **A** and of −16.1% for the measurement shown in **B**.

End-expiratory lung volume, end-tidal concentration measurement, and unusual rises in expired N2 concentration (leaks, trapped gas) were the tests which most often predicted the largest effect, and were most frequently above the threshold of an estimated impact on LCI of 15% (Table 2).

**Table 2:** Individual effects of quality control tests for children with CF (dataset 1), from 2471 measurements. The table shows how often each test showed a result that exceeded the acceptability threshold of 15%, as well as how often it was the largest source of error for the measurement. ET: End of test, EELV: End-expiratory lung volume, V_T_: Tidal volume.

| EFFECT NAME | EXCEEDS THRESHOLD [%] | LARGEST ERROR [%] |
| --- | --- | --- |
| LEAK, TRAPPED GAS | 9.6 | 30.7 |
| ET CONCENTR. METHOD | 4.3 | 19.2 |
| EELV VARIATION | 3.8 | 22.9 |
| SIGNAL RESYNC | 2.3 | 15.9 |
| V<sub>T</sub> SIZE | 1.7 | 5.9 |
| BREATH DETECTION | 0.7 | 1.6 |
| V<sub>T</sub> VARIATION | 0.2 | 3.8 |
| OVERFLOW | 0.1 | 0.0 |

The overall QC acceptance ratio (the proportion of visits which had no test which had a higher estimated effect than 15%) was 76%. The mean per-visit variance of LCI was 0.72 TO when including all measurements, and reduced to 0.39 TO when only including measurements accepted by the automated QC algorithm.

The QC acceptance ratio per age group is shown in supplementary table S1, confirming that QC-related outcomes improve as the children age and become more adept at following the instructions around a MBW measurement.

### Association to LCI

There is a significant correlation of the within-visit variation of the lung clearance index (LCI) and the impact estimate by the quality control algorithm. The combined estimate of all quality control tests for the impact on LCI managed to explain a significant proportion of the within-visit variability between individual MBW tests (R^2^ = 0.45) (Figure 3 and Table 3).

**Figure 3:**
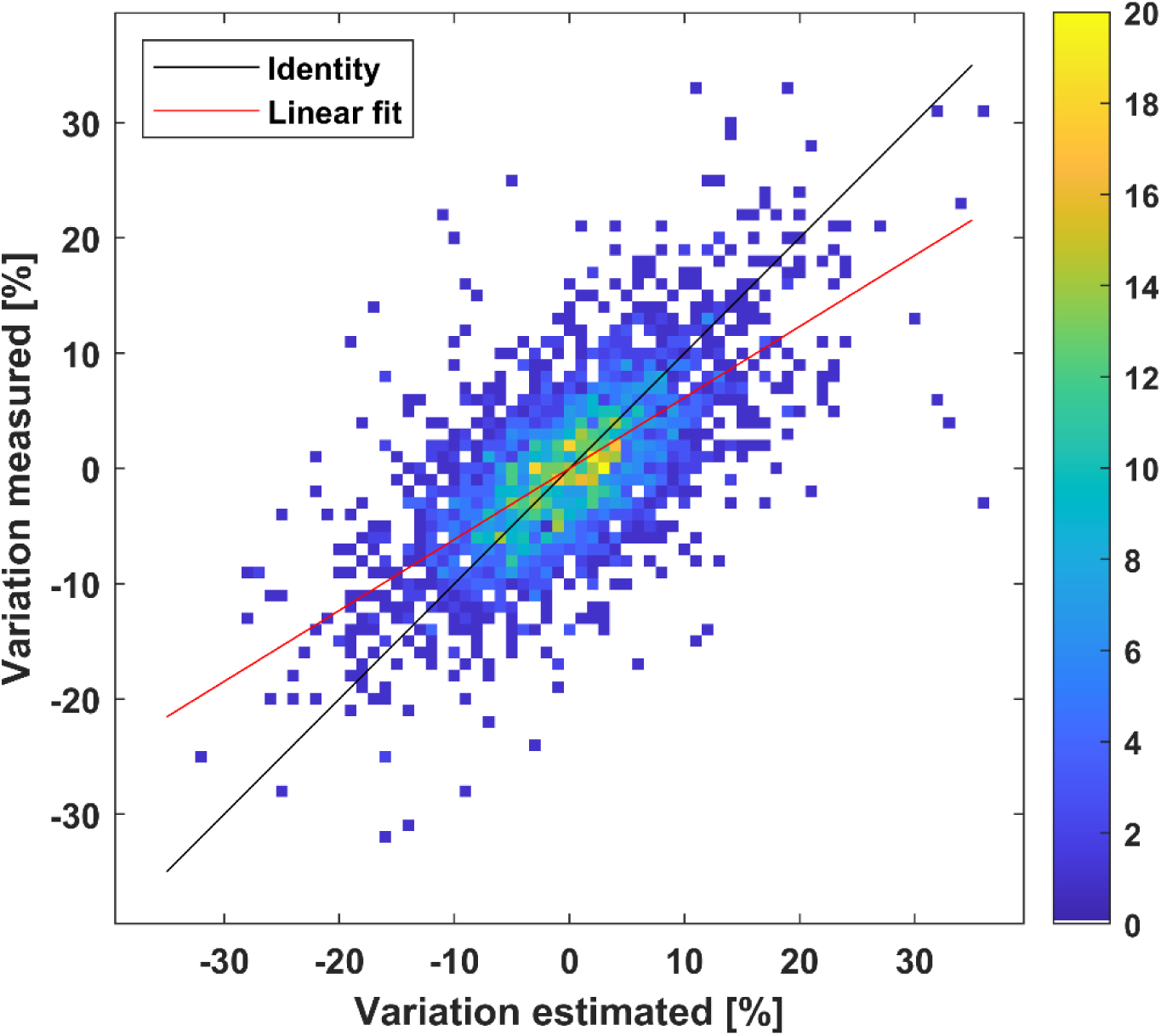
Comparison of aggregated estimate of impact of QC-related factors (x-axis) vs measured within-visit variation of LCI values (y-axis) relative to the visit mean in trials in children with CF (dataset 1). R^2^=0.45. The scatterplot is represented as a heatmap, where color intensity represents the number of data points within a 1% by 1% range. The line of identity shows where the data points would be situated if the automated QC algorithm managed to explain all within-visit variability, the red line shows the linear fit.

**Table 3:** R^2^ of the relation of either individual or combined estimated effects on LCI with measured within-visit variability of LCI in a dataset of 2471 multiple-breath washout measurements in children with cystic fibrosis. ET: End of test, EELV: End-expiratory lung volume, V_T_: Tidal volume.

| EFFECT NAME | R<sup>2</sup>(%) |
| --- | --- |
| COMBINED | 45.0 |
| EELV VARIATION | 18.7 |
| ET CONCENTR. METHOD | 10.4 |
| LEAK, TRAPPED GAS | 5.8 |
| V<sub>T</sub> SIZE | 3.9 |
| SIGNAL RESYNC | 1.9 |
| BREATH DETECTION | 1.6 |
| OVERFLOW | 0.5 |
| V<sub>T</sub> VARIATION | 0.3 |

The individual tests did not all contribute equally to the correlation with within-visit variability, with the largest share of the correlation explained by the effect of variable end-expiratory lung volume (EELV), followed by end-of-test determination via end-tidal concentration, leaks and release of trapped gas, and tidal volume size, in that order. Finally, the correlation of the technical issues related to synchronization, breath detection, and inspiratory overflow leaks, as well as tidal volume variability with within-visit variability was minor (Table 3).

The association of measurements of individual visits are shown in supplementary figure S3. The median Pearson correlation coefficient of within-visit LCI with estimated effects of QC-related factors was 0.83 (Supplementary Figure S4).

### Relation to expert rating

The agreement between the automated QC and expert QC was only moderate, with a Cohen’s kappa ranging from 0.32 between the automated QC and the expert rating on the day of the test, to 0.46 between the automated QC and the retrospective expert QC. The agreement between the two instances of expert QC was higher with a Cohen’s kappa of 0.65 (Supplementary Table S2).

The association of the critical variable for automated QC (highest results of estimated impact over all tests) with the grading from expert QC is shown in Figure 4a. Figures 4b, c illustrate the disagreement between automated and expert QC for two measurements.

**Figure 4:**
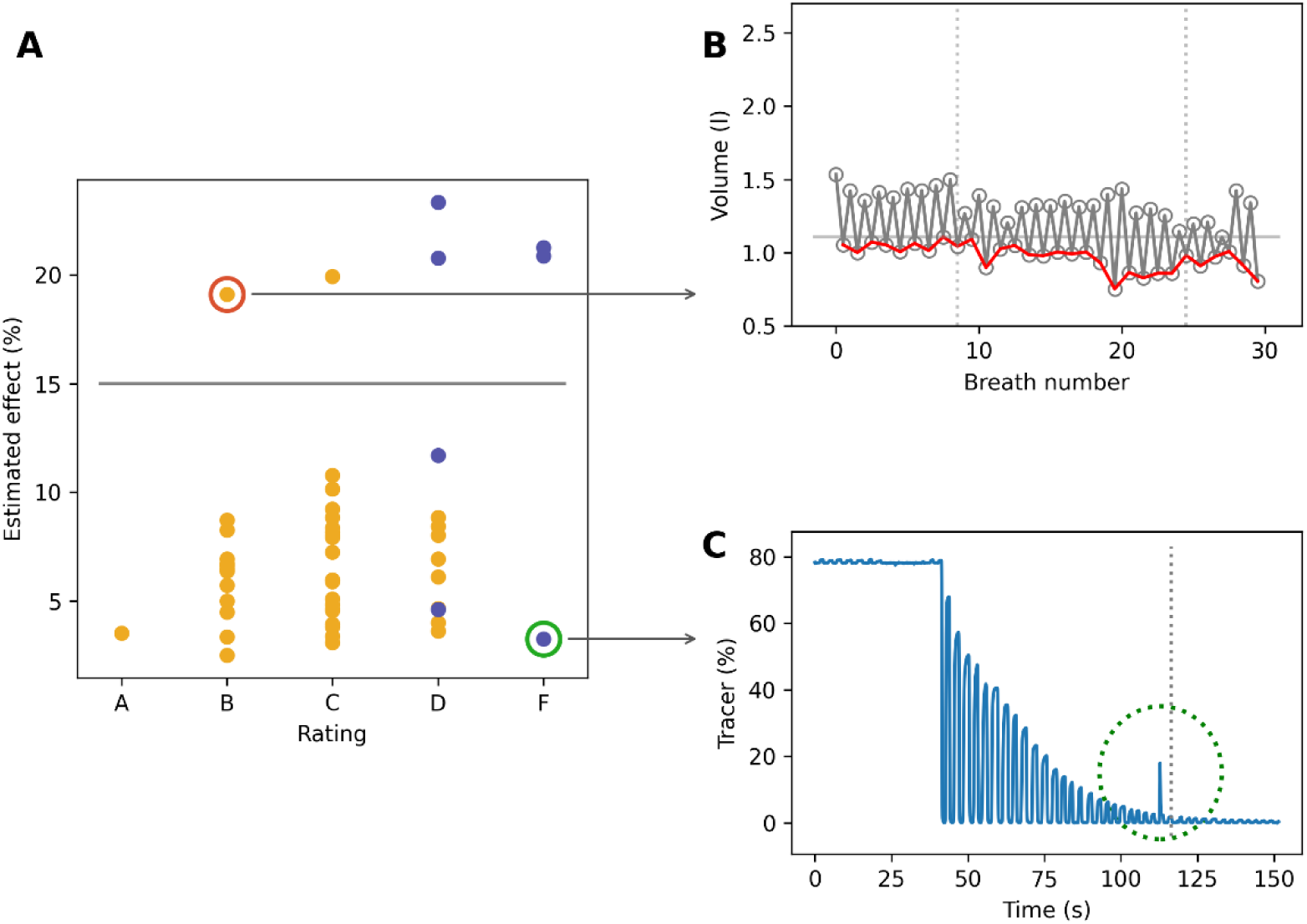
Estimated impact of automated QC versus expert QC. **A** shows the highest automated QC test result of a measurement versus the retrospective expert QC ratings (A-F) of one expert for the 50 measurements of healthy children (HC) in dataset 2. Each dot corresponds to one MBW measurement, accepted/rejected by expert rater (orange/blue). For the red-circled sample, changes in end-expiratory lung volume (EELV) result in an estimated impact of almost 20% in LCI. **B** shows the corresponding lung volume of this measurement (grey line) together with the EELV (red line). The dotted lines show the washout start and end. **C** shows the tracer concentration for the green circled measurement in A. A leak (marked by dotted circle) during expiration towards washout end (dotted line) is clearly visible, resulting in an expert QC F-rating. The volume of tracer gas corresponding to the leak is small, and visually, the end-expiratory tracer concentration follows the expected exponential decrease, unaffected by the leak. Computation indicates that the impact of this leak on LCI and FRC is below 5%.

## Discussion

### Summary

We developed a novel, fully automated quality control (QC) tool for multiple-breath washout (MBW) measurements, which gauges the relevance of behavioral irregularities by estimating the direct impact of those irregularities on MBW outcomes. We applied this algorithm to datasets of healthy children and children with cystic fibrosis (CF), and showed that it could explain a significant part of existing within-visit variability in MBW outcomes. Further, we characterized the differences between it and the current gold standard expert QC. The development of a fast, automated, reproducible, and standardized approach to MBW QC represents a significant advance for MBW as a measurement technique.

### Discussion of results

The automated QC approach described here provides the first estimate of the impact of individual quality control issues on MBW outcomes. This allows for rapid identification of problematic behaviors within MBW measurements, and allows the relevance of different sources of error to be judged according to the same comparable scale, namely impact on MBW outcomes.

The examples shown in figure 2 illustrate the potential difficulty to visually judge the impact of factors such as variable end-expiratory lung volumes (EELV) on MBW outcomes, as they can be transient, quickly reverse, or have different magnitudes of impact when they occur early or late in the measurement. The model-based approach used here can quantify the expected impact of these behavioral irregularities in a reproducible manner across measurements.

We could show that a significant part of the within-visit variability of MBW measurements in a large dataset of children with CF was due to factors linked to QC that could be estimated using this new algorithm. The highest correlated individual factors within this aggregated estimate were changing EELV and the breath-size-dependent determination of end-expiratory N_2_ concentration. The QC-related factor which most frequently exceeded the threshold of acceptability in a dataset of children with CF was trapped gas and leaks, related to end-expiratory N_2_ concentrations that deviated from washout-model-based expectations. This is likely due to MBW measurements in children with CF often showing extended washout “tails”, where the end-tidal concentration is slowly dropping below the end-of-test threshold, but then occasionally slightly increases again as trapped gas is released. While we used a fixed 15% as the threshold of acceptability for individual tests here, it might make sense in certain cases to instead relate the estimated impact to clinically relevant thresholds. E.g. if an effect of >15% is estimated due to unexpected washout progression, the LCI might be so clearly above the upper limit of normal that the estimate does not change the clinical outcome of the test. In general, an advantage of the approach described here is that QC can be automatically performed according to the needs of the study or use case. Some applications might have different objectives that require more or less strict QC based on the individuals at hand, or ignore certain specific tests according to the question.

### Comparison to expert QC

We could show that the results of MBW QC showed moderate overlap to those obtained by gold standard expert QC. While the overlap between automated and expert QC, especially within the group of rejected measurements, was not perfect, this overlap was not much lower than between two separate instances of expert QC. This is likely due to several key differences between the approach described here and the current expert QC gold standard. The current standard e.g. states that any leak (no matter the size) should lead to exclusion of the measurement. Similar to how other breathing artifacts exist on a «spectrum of acceptability», the automated QC allows for small potential leaks, as long as the estimated impact on MBW outcomes remains small. Additionally, the current gold standard defines limits of acceptable deviation from the ideal behavior for each QC test individually, while in the automated QC they are replaced by the same scale based on the impact on main MBW outcomes. Figure 4 shows two examples of measurements where the automated QC and current gold standards disagree in their final assessment of acceptability. In one case, expert QC may have struggled to visually identify whether an apparent change in EELV was sufficient to influence outcomes. In the other, there is clearly visible evidence of a small leak, which expert QC is instructed to reject, but which the automated QC identified as having little effect on outcomes.

Clear advantages of automated QC are that it saves time, provides a single, repeatable standard for QC across data from different centers, and that it can be retrospectively applied to data when it is re-analyzed according to the newest MBW algorithms. Furthermore, a major difficulty in expert QC is to determine whether an anomalous signal (such as an uptick in end-tidal N_2_) is due to a severe QC factor such as a leak, or whether the signals can be entirely explained by the underlying breathing behavior. The model based approach used here inherently distinguishes between these cases, which is particularly useful in cases where the test subject is unable to attain a high quality of tidal breathing.

### Comparison with other studies

At the core of the approach described here lies the washout model. In a previous study, a simpler model of homogeneously ventilated lungs using constant volumes was employed to quantify the impact of the size of tidal volume relative to FRC on LCI ^21^. The more general model described here allows to quantify irregular breathing behavior, e.g. changes in EELV, which have a major effect on measured LCI variations. Additionally, it models ventilation efficiency.

A two compartment model of washout using a superposition of two linear components was described in a previous study^16^, which allows for an analysis of slow and fast ventilating compartments in MBW. The use of a linearly decreasing washout efficiency as a function of washout progress used here is a similar concept (leading to a decreasing speed of washout as the washout progresses), though the precise implementation differs.

Previous research has demonstrated that suboptimal data processing by proprietary MBW device software may occur^22, 23^, potentially affecting the accuracy of measured LCI values^17, 24–26^. Included in the automated algorithm are therefore alternate methods for signal synchronization, breath detection, and end-tidal concentration calculation. These are not strictly related to detecting irregular behaviors of the test subject, but instead aim to detect behaviors where the choice of processing methodology might have a major impact.

### Strengths and limitations

We present a novel framework for interpreting MBW and performing quality control in an automated and objective manner. We examined the use of these new tools in a large dataset of children with cystic fibrosis (CF), and compared its application to expert QC in a dataset of heathy controls and children with CF.

The algorithm described here was developed for and applied to N_2_MBW data measured with the Ecomedics Exhalyzer^®^ D. Specifically, the automated QC relies on the presence of a CO_2_ signal, a tracer gas signal, and a flow signal. Some tests, such as the synchronization and inspiratory overflow tests, are specific to the Exhalyzer^®^ D device setup. However, only minor adaptations would be required in order to extend the algorithm to SF_6_MBW measurements, and, depending on the device setups, measurements originating from other devices.

A desirable time-saving feature of MBW QC would be live feedback for people as they perform the test, informing them when a behavior is likely to have compromised a test and a restart would be advisable. With the current implementation of the automated QC, the earliest time to perform the QC is at the end of the measurement. Further development would be needed to make automated QC during the measurement possible.

## Conclusion

This work introduces a novel, innovative approach to automated quality control for MBW. It demonstrates a feasible method to assign estimates of impact to behavioral irregularities in MBW measurements in a fast, objective and reproducible manner, which permits fully automated QC but especially facilitates the QC of measurements with intermediate quality, which are difficult to rate by visual inspection. Furthermore, it provides novel tools with which to interpret MBW, and provides new insights into the thresholds of acceptability of MBW measurements.

## Supporting information

Supplement_prerpint

## Acknowledgements

We thank our study participants and their families for allowing their data to be used for research, and we thank the study nurses and clinicians for their work in conducting the measurements.

## Author contributions

FW developed the QC algorithm, FW and SB (re-)analyzed the data and wrote the manuscript. FW and MC developed the framework of washout efficiency and the related ventilation inhomogeneity outcomes. MC edited the manuscript. XB co-developed the QC algorithm and edited the manuscript. BF provided the existing longitudinal CF dataset. PL co-conceived of the research and edited the manuscript.

## Data availability statement

The data that support the findings of this study are available on request from the corresponding author. The data are not publicly available due to privacy or ethical restrictions.

