## Supplement_prerpint for "Automated and impact-based quality control in multiple-breath washout"

### Online Supplement

#### 1 Methods

##### 1.1 Washout efficiency

According to the washout model in equations (1) and (2), on a logarithmic scale, for each breath the washout efficiency is given by the washout progress (measured reduction in $N_2$ concentration) relative to *log(K)*,

$$f = \frac{\log(N_{2-new}) - \log(N_2)}{\log(K)} \qquad \text{(S1)}$$

where

$$K = \frac{V_A + V_D}{V_A + V_{insp}}$$

with $V_A$ being the alveolar volume, $V_D$ the dead space volume, and $V_{insp}$ the inspired volume of a given breath.

For homogeneously ventilated lungs (by definition f=1) the washout progress is equal to $\log(K)$. Thus, equation (S1) states that the washout efficiency is equal to the washout progress relative to the washout progress in homogeneously ventilated lungs.

Calculating the normalized logarithmic concentration of nitrogen ($N_2$) allowed for the calculation of washout process in the range [0, 1]. The normalized logarithmic concentration was calculated by the logarithm of the normalized end-tidal concentration of $N_2$ relative to the initial concentration $N_{2-initial}$ divided by the logarithm of the normalized end-of-test concentration of $N_{2-final}$ (0.025 = 2.5%) relative to the initial concentration,

$$N_{2-lognorm} = \frac{\log\left(\frac{N_2}{N_{2-initial}}\right)}{\log\left(\frac{N_{2-final}}{N_{2-initial}}\right)} \qquad \text{(S2)}$$

#### 1.2 Estimating washout efficiency

The washout efficiency f was modelled as a linear function of normalized logarithmic $N_2$ concentration. Measured efficiency was calculated according to equation (S1) for every breath and used to estimate intercept and slope using a robust procedure. This is illustrated in figure S1 and more details about the robust procedure are explained in the figure caption.

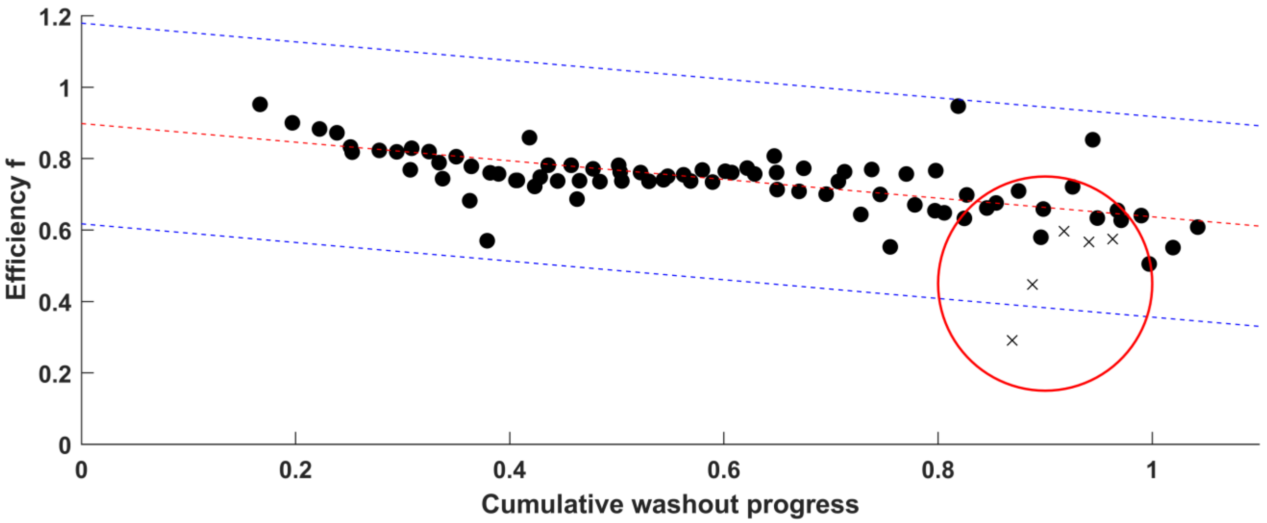


**Supplementary figure S1:** Illustration of washout efficiency parameters. Black dots show the calculated efficiency for breath pairs as a function of linearized cumulative washout progress (0: initial concentration, 1: $N_2$=2.5%). The red dotted line represents a linear best fit. To improve accuracy of the fit, each breath generates five data points corresponding to the dilutions between it and the next five breaths. Any breath where one of these dilution efficiencies falls outside of the fit ± 3 x the median absolute deviation from the fit ± a base value of 0.2 (shown by the blue dashed lines), is excluded (shown by the group of five crosses circled in red, representing one breath (five breath-pairs) excluded from the fit).

#### 1.3 Most efficient $V_T$

In order to judge the effect of the tidal volume size $V_T$ chosen by the test subject, we compared the results obtained in simulations using constant inspired and expired volumes, at either the measured median $V_T$, or the $V_T$ that is calculated to be most efficient. The most efficient $V_T$ was defined as the one which would likely produce the smallest lung clearance index (LCI), and therefore had the lowest required cumulative expired volume (CEV). At low volumes, the dead space interferes with the dilution of $N_2$, making most of the volume exhaled useless at washing out the lungs. At large volumes, the dilutions become less volume efficient, as multiple small dilutions of the same volume as one large dilution are more efficient. To find the most efficient $V_T$ we therefore calculated the washout progress $P_{breath}(V_T)$ for a range of possible values of $V_T$. We calculated the expected number of breaths of that volume required to wash out the lungs as

$$n = 1/P_{breath}(V_T).$$

The most efficient $V_T$ was then the one that minimized $CEV \approx n * V_T$ , figure S2.

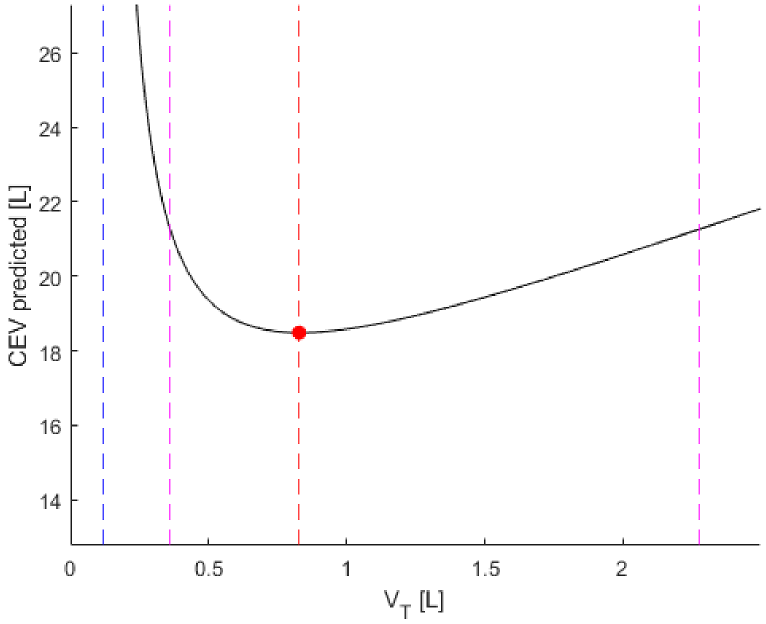


**Supplementary figure S2:** CEV predicted by the washout model as a function of $V_T$ for an example measurement. As $V_T$ increases beyond the minimum threshold of the total dead space (blue dashed line) washout becomes more volume efficient, until it reaches the ideal $V_T$ (red dashed line). The purple dashed lines indicate where the CEV is expected to increase more than 15% due to $V_T$ size.

#### 1.4 End-tidal concentration measurement:

In the Spiroware 3.3.1 algorithm for the analysis of MBW measurements, the end-tidal tracer concentration measurement depends on the size of breath, as the calculation is based on the average of the $N_2$ signal in the 90-95%-window of expired volume of a given breath. We developed an alternative method for the determination of end-tidal concentration, which was designed to be less sensitive to changes in breath size. For this, we first calculated the median $CO_2$ signal as a function of expired volume. We then used this to determine the best linear fit of a given breath's $N_2$ signal to this average $CO_2$ signal. As both these signals shared a similar shape, we could determine a linear fit of $N_2$ to average $CO_2$. By evaluating the linear fit at the median $CO_2$ concentration corresponding to the median expired volume, we were able to obtain an estimate of $N_2$ concentration which was less dependent on breath size.

Re-calculating MBW results using this method of end-tidal concentration calculation provided an estimate of where the MBW measurement would have ended had the breath sizes around end-of-test been equal to the median expired volume.

#### 1.5 Signal resynchronization:

Signal synchronization, i.e. temporal alignment of the measured $O_2$ and $CO_2$ signals, is important for accurate calculation of $N_2$ concentration. Signal synchronization was performed using the mean delay times estimated by three separate methods: i) analogous to the Exhalyzer® D calibration, but based not on a separate calibration but on the signals of the measurement, ii) maximizing cross-correlation between the $O_2$ and CO2 signals, and iii) minimizing signal noise as estimated by the sum of the absolute values of the first derivative of the N2 signal.

Re-calculating MBW results of a measurement with recalibrated delay values provided an estimate of whether the calibration was good enough for it not to have a negative effect on MBW outcome accuracy. If there was a large difference in MBW outcomes due to a re-analysis with calibrated delay times, then the delay times recorded in the A-file-header were no longer correct at time of measurement, or were not compatible with a change in methodology (e.g. delay times recorded before switching to Spiroware 3.2.1, which introduced dynamic delay correction, which needs different calibration values).

### 2 Results

#### 2.1 Objective QC

| AGE RANGE | NUMBER OF MEASUREMENTS | QC ACCEPTANCE RATIO (%) |
|---|---|---|
| **(3, 6]** | 184 | 61 |
| **(6, 8]** | 489 | 71 |
| **(8, 10]** | 454 | 77 |
| **(10, 12]** | 415 | 82 |
| **(12, 14]** | 337 | 83 |
| **(14, 16]** | 287 | 72 |
| **(16, 19]** | 305 | 80 |
| **OVERALL** | 2471 | 76 |

**Table S1:** Measurement acceptance ratio per age group according to QC criteria, from 2471 measurements of children with cystic fibrosis.

#### 2.2 Association to LCI

Measurements of four randomly selected visits are show in figure S2 with 8, 3, 4, and 4 measurements, respectively.

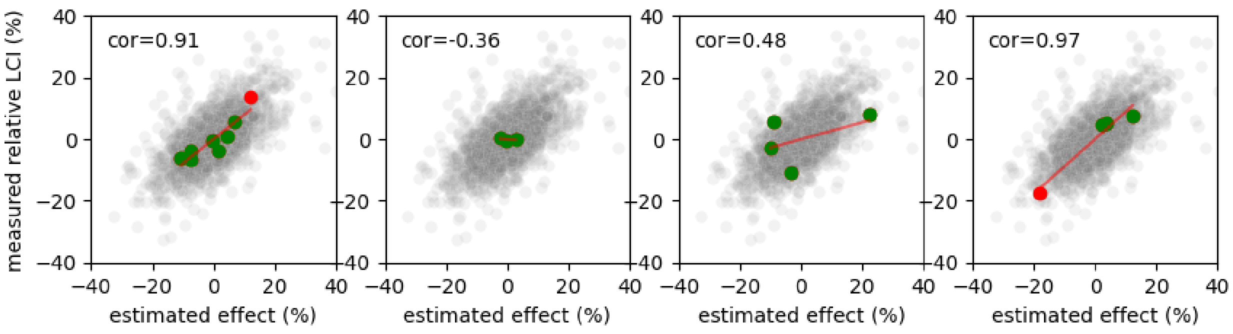

**Figure S3:** Measured relative LCI versus relative estimated effect of all measurements in light grey as in figure 3. Each subplot shows samples of one visit (green if accepted by QC, red otherwise) with Pearson correlation and a linear fit (red line) for illustration. Estimated effect is the aggregated effect. Measurements are accepted by QC if all individual effect are smaller than 15%, though the estimated aggregated effect can be larger.

The correlation of the measurements in each visit shown in figure S3 is shown in the top left corner. The histogram of these correlation coefficients for all visits is shown in figure S4.

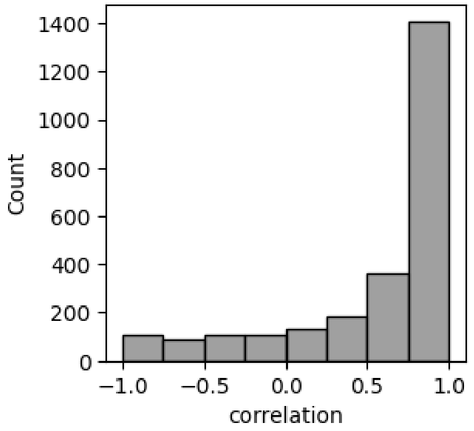


**Figure S4:** Histogram of per-visit correlation of aggregated estimated effect by the automated QC algorithm with measured LCI. A correlation of 0 indicates that the estimated effect does not correlate with within-visit differences in LCI. A correlation of 1 indicates that all within-visit difference is proportional to the estimated effect, though the magnitude of the impact estimate could still be incorrect. Median correlation for all visits is 0.83.

#### 2.3 Relation to expert rating

| | | Expert QC 1: 1 | Expert QC 1: 0 |
|---|---|---|---|
| Auto QC | 1 | **80** | **9** |
| | 0 | **6** | **5** |

| | | Expert QC 2: 1 | Expert QC 2: 0 |
|---|---|---|---|
| Auto QC | 1 | **83** | **6** |
| | 0 | **5** | **6** |

| | | Expert QC 2: 1 | Expert QC 2: 0 |
|---|---|---|---|
| Expert QC 1 | 1 | **83** | **3** |
| | 0 | **5** | **9** |

| | Auto QC vs Expert QC 1 | Auto QC vs Expert QC 2 | Expert QC 1 vs Expert QC 2 |
|---|---|---|---|
| Agreement ($p_0$) | 85.0 % | 89.0 % | 92.0 % |
| Random agreement ($p_e$) | 78.1 % | 79.6 % | 77.4 % |
| Cohen's κ | **0.32** | **0.46** | **0.65** |

**Table S2: Agreement between automated and expert QC.** Shown is the number of measurements which were considered acceptable (1) or rejected (0) by either the automated QC (Auto QC), the expert rater on the day of the original measurement (Expert QC 1) or the retrospective QC by an expert rater (Expert QC 2). Additionally shown is the agreement between the instances of QC, the probability of random agreement, and the resulting Cohen's kappa.

### 3 Source code

Full details of the QC algorithm can be found in the source code of the implementation in MATLAB. The source code can be found on GitHub,

https://github.com/silviobe/mbw_quality_control.git

and the README files of the project gives an overview of the implementation.

Note, for technical reasons, breath detection and signal re-synchronization is performed in a separate software package, analogous to Spiroware 3.3.1, and is not included in the source code.
